# Sleep Quality Mediates the Relationship Between Sleep Hygiene and Psychological Stress Among Adults With Multiple Cardiovascular Risk Factors

**DOI:** 10.1101/2024.11.25.24317945

**Authors:** Xiaoyue Liu, Junxin Li, Jinyu Hu, Jason Fletcher, Yvonne Commodore-Mensah, Cheryl R. Himmelfarb

**Affiliations:** New York University Rory Meyers College of Nursing; Johns Hopkins School of Nursing

**Author notes:** **Corresponding Author** Xiaoyue Liu, PhD, RN, 433 1^1920^ Ave., New York, NY, 10010.

**Keywords:** Home environment, Bedtime behaviors, Sleep quality, Psychological stress, Cardiovascular risk factors

## Abstract

**Background:** Poor sleep quality and psychological stress are interrelated and disproportionately affect adults with multiple cardiovascular disease (CVD) risk factors. Maintaining an optimal home environment and engaging in healthy bedtime behaviors are important components of sleep hygiene practices that influence sleep health and mental well-being. However, research is scarce in exploring the associations between sleep hygiene, sleep quality, and psychological stress among adults with multiple CVD risk factors.

**Methods:** A cross-sectional study was conducted with 300 adults diagnosed with hypertension and type 2 diabetes. Individuals were recruited from a large academic health center and were asked to complete an online survey. Sleep hygiene was assessed by nine individual factors focusing on negative household environment (safety, physical comfort, temperature, and light) and poor bedtime behaviors (watching TV, playing video games, using small screens, and eating) and by a composite score. Multiple regression and mediation analysis with bootstrapping were employed to examine the associations.

**Results:** Of the sample, 78% reported poor sleep quality and 44% reported high psychological stress. Individual sleep hygiene factors as well as the composite score were significantly associated with poorer sleep quality and higher psychological stress. Sleep quality mediated the association between the composite score and psychological stress (Indirect effect: 0.16; 95% bootstrap confidence interval: 0.04-0.35).

**Conclusions:** The study highlights strong links between sleep hygiene, sleep quality, and psychological stress. Although causality cannot be established, current evidence suggests that promoting home environment and bedtime behaviors may alleviate sleep and psychological burdens in adults with multiple CVD risk factors.

**CLINICAL PERSPECTIVE:** *What Is New?:*

- Adults with hypertension and type 2 diabetes often experience poor sleep quality and elevated psychological stress.
- Room environment and bedtime behaviors are important components of sleep hygiene practices that strongly associate with sleep quality and mental well-being.
- Sleep quality may serve as a mediating factor between sleep hygiene and psychological stress.

*What Are the Clinical Implications?:*

- Enhancing room environment and bedtime behaviors is crucial for adults with multiple cardiovascular disease risk factors.
- Given the crucial roles of both sleep and psychological stress in cardiovascular health, promoting modifiable sleep hygiene practices may be an effective strategy to mitigate health risks in this population.

## INTRODUCTION

Cardiovascular disease (CVD) remains the leading cause of death in the U.S.^1^ Hypertension and type 2 diabetes are two of the most common risk factors of CVD, affecting 122.4 million and 29.3 million U.S. adults, respectively.^1^ These chronic conditions frequently co-exist. Adults who have hypertension for 5–10 years are four times more likely to develop diabetes compared to those with hypertension for less than five years.^2^ Similarly, up to 75% of adults with diabetes have hypertension.^3^ Together, people with both conditions are at substantially increased risks of CVD morbidities, such as stroke, coronary heart disease, congestive heart failure, and peripheral vascular disease.^4^

Psychological stress is a critical risk factor for CVD.^5–9^ A large population-based cohort study conducted across 21 countries found that higher levels of stress were associated with increased risks of developing CVD (hazard ratio [HR]: 1.22, 95% confidence interval [CI]: 1.08-1.37) and stroke (HR: 1.30, 95%CI: 1.09-1.56) over a median follow-up of 10 years.^10^ Psychological stress prevalently affects individuals with multiple CVD risk factors. One study revealed that nearly half of participants who had hypertension and/or diabetes suffered from psychological stress.^11^ While several methods have been explored to mitigate stress, growing evidence suggests that improving sleep may be an effective strategy to alleviate mental distress. Clinical trials indicate that sleep promotion interventions lead to significant small-to-medium-sized effects in reducing psychological stress.^12^ This evidence is particularly relevant to individuals at risk for CVD, as better sleep can not only reduce stress but also improve cardiovascular health.

Room environment and bedtime behaviors are essential components of sleep hygiene practices. While their effects on sleep are well-established, many factors within these components may also influence psychological stress. For example, living in an unsafe household can serve as a direct source of daily stress.^13^ Additionally, contextual factors such as uncomfortable temperature, excessive noise, and ambient light exposure can disrupt sleep.^14^ Chronic sleep disturbances may cause dysregulation of emotional responses and activation of the hypothalamic-pituitary-adrenal axis, which in turn exacerbates stress responses.^15,16^ Beyond the living environment, research indicates that nighttime behaviors may contribute to poor sleep. Johnson and colleagues^17^ found that listening to the radio or music, reading books, and consuming meals or snacks in-bed were significantly related to actigraphy-based sleep measures in a cohort of African-American older adults. Although there is limited research linking bedtime behaviors to psychological stress, it is plausible that these activities may indirectly raise stress levels through poor sleep.

Currently, few studies have examined sleep hygiene practices, specifically room environment and bedtime behaviors, in adults with multiple CVD risk factors. To address this, we aimed to investigate this topic and expand upon existing knowledge by exploring the associations between sleep hygiene, sleep quality, and psychological stress. We hypothesized that sleep quality would be independently associated with sleep hygiene and psychological stress and that sleep quality would mediate the association between sleep hygiene with psychological stress.

## METHODS

### Study Design and Setting

We conducted a cross-sectional study among adults who received primary care at a large academic healthcare system in the Maryland and Washington D.C. metropolitan areas.

### Participants

Participants were identified through a retrospective review of electronic health records. The inclusion criteria were: 1) aged 18 years or older; 2) had a diagnosis of primary hypertension (I-10) either from a recent encounter or listed as an active problem within the last 12 months; 3) had a diagnosis of diabetes (I-11) either from a recent encounter or listed as an active problem within the last 12 months; 4) had the most recent systolic blood pressure ≥ 130 mm Hg and HbA1c ≥ 6.5%. The exclusion criteria were: 1) treated as inpatients; 2) pregnant or lactating; or 3) had conditions including type 1 diabetes, gestational diabetes, end-stage renal disease, acute cardiovascular event, acute cerebrovascular event, or metastatic cancer.

### Data Collection

Each participant completed an anonymous online survey, which included questions on sociodemographic backgrounds, clinical characteristics, room environment, bedtime behaviors, sleep quality, and psychological stress. This study was approved by the university’s Institutional Review Board.

### Variables

#### Psychological Stress

was measured by the Perceived Stress Scale (PSS).^18^ The 4-item instrument examines the level at which situations are appraised as stressful in an individual’s life. The PSS-4 estimates how unpredictable, uncontrollable, and overloaded individuals perceive their lives to be. Respondents rate the occurrence of their feelings or thoughts in the past month on a scale of 0 (never) to 4 (very often). The total score is calculated by summing all ratings across the items, with an average score ≥ 6 indicating high levels of stress.^19^ The PSS-4 has been considered a valid tool, with adequate reliability, for assessing psychological stress.^20,21^

#### Sleep Quality

was measured by the Pittsburg Sleep Quality Index (PSQI).^22^ The 19-item instrument evaluates sleep quality and disturbances over the past month. The items cover seven components, including subjective sleep quality, sleep latency, sleep duration, habitual sleep efficiency, sleep disturbances, use of sleep medication, and daytime dysfunction. These components are summed to generate a global score ranging from 0 to 21, where higher scores reflect poorer sleep quality. A global PSQI score greater than 5 indicates poor sleep quality. The PSQI has demonstrated satisfactory internal consistency, reliability, and construct validity across diverse populations.^23–25^

#### Sleep Hygiene (home environment and bedtime behaviors)

was measured using the questions developed by Johnson and colleagues.^17^ We created an adapted version by including the questions that adversely impact sleep. The adapted instrument consisted of 9 items, with 5 assessing household environment and 4 assessing bedtime behaviors. The household environment questions assessed the following aspects: unsafe household, uncomfortable physical environment, uncomfortable temperature, noise disturbances, and light disturbances. The bedtime behavior items included watching TV, using small screens (e.g., computer, tablet, or phone), playing video games, and eating meals or snacks. Following the scoring approach suggested by the authors, all responses were coded on a scale of 0 to 1 (See Supplemental Table 1). A composite score was then created based on the average of 9 items described above (range 0-1), where higher scores represented poorer room environment and bedtime behaviors. To facilitate interpretation, the composite score and four components with more than two responses (unsafe household, uncomfortable physical environment, uncomfortable temperature, noise disturbances) were standardized to a normal distribution with a mean of 0 and a standard deviation of 1.

#### Covariates

Age and body mass index (BMI) were assessed as continuous variables. Other sociodemographic variables were categorized as biological sex (male or female), race (White or non-White), education (high school/general educational development [GED] or less, associate degree/some college, and bachelor’s degree or above), employment status (employed or unemployed), marital status (married or not married), home ownership status (yes or no), and history of obstructive sleep apnea (OSA) (yes or no).

### Statistical Methods

Descriptive statistics were reported as mean ± standard deviation for continuous variables and frequency (%) for categorical variables. Multiple linear regression was used to examine the individual component and the composite score of sleep hygiene with sleep quality as well as psychological stress. The models were adjusted for sex, age, race, BMI, marital status, employment, home ownership, and history of OSA. Mediation analysis was performed to examine whether sleep quality mediated the association between the composite score of sleep hygiene and psychological stress. To assess mediation, four pathways were estimated (Figure 1): <u>path a</u> – the effect of the home environment and bedtime behaviors composite score (independent variable) on sleep quality (mediator), <u>path b</u> – the effect of sleep quality (mediator) on psychological stress (dependent variable), adjusting for the composite score, <u>path c</u> – the total effect of the composite score on psychological stress, and <u>path c’</u> – the direct effect of the composite score on psychological stress after accounting for sleep quality. The indirect effect is estimated as the product of the a- and b-pathways (a*b) and represents the mediated effect. Mediation is identified when the direct effect is less than the total effect (c’ < c). It is considered full mediation when c’ = 0 and partial mediation when c’ ≠ 0. Bias-corrected accelerated (BCa) bootstrap confidence intervals (CI) with 5,000 resamples were generated for estimates of direct, indirect, and total effects.^26^ The percentage of the total effect that is mediated was calculated by dividing the indirect effect by the total effect (% mediated = a*b/c).^26^ All analyses were performed using R version 4.3.1.^27^ The statistical significance was set at *p* < 0.05.

**Figure 1.**
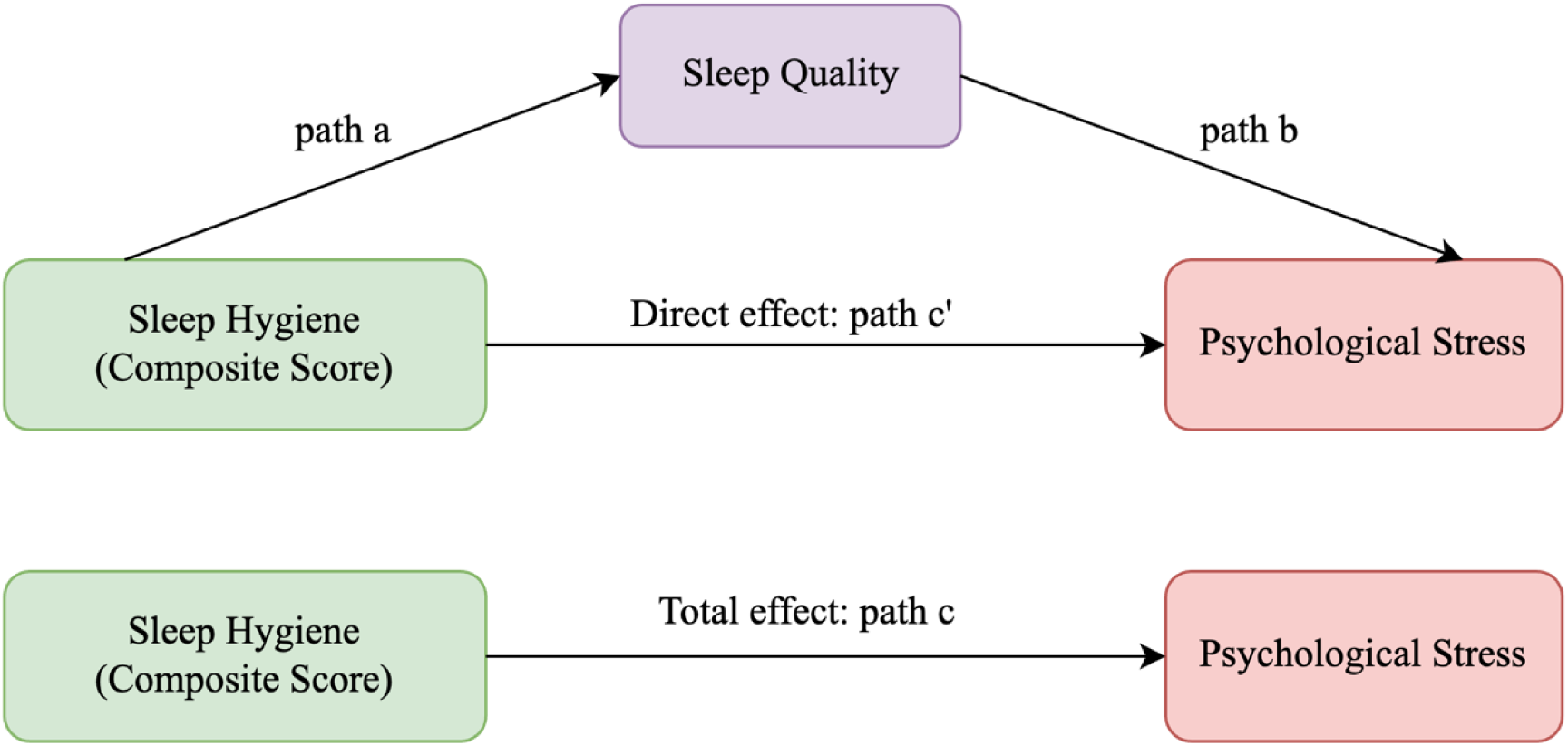
Mediation analysis of the composite score of sleep hygiene, sleep quality, and psychological stress.

## RESULTS

### Characteristics of Study Participants

Three hundred adults diagnosed with hypertension and type 2 diabetes participated in the study. The mean age was 62 ± 12 years. Over half were female, 58% were White adults, 72% reported home ownership, and 67% had a history of OSA. Of the sample, about 78% experienced poor quality of sleep in the past month, and 44% of the sample reported a high level of psychological stress. The practices of unhealthy sleep hygiene ranged from 8% for uncomfortable bedroom temperatures to 66% for using small screens at bedtime (Table 1).

**Table 1.** Characteristics of Participants (N=300)

| **Study Variables** | **N (%)** |
| --- | --- |
| **Sociodemographic Characteristics** |  |
| Age, mean ± SD | 62 ± 12 |
| Sex |  |
| Male | 146 (49%) |
| Female | 152 (51%) |
| Race |  |
| White | 174 (58%) |
| Non-White | 126 (42%) |
| Education |  |
| High school/GED or less | 43 (14%) |
| Associate degree/some college | 100 (34%) |
| Bachelor's degree or above | 154 (52%) |
| Employed | 153 (51%) |
| Married | 173 (58%) |
| Home ownership | 215 (72%) |
| **Clinical Characteristics** |  |
| Body mass index, mean ± SD | 33.4 ± 7.9 |
| Sleep quality (Pittsburgh Sleep Quality Index score), mean ± SD | 7.9 ± 3.9 |
| Good sleeper | 65 (22%) |
| Poor sleeper | 235 (78%) |
| Psychological Stress (Perceived Stress Scale score), mean ± SD | 5.0 ± 3.3 |
| Low stress | 169 (56%) |
| High stress | 131 (44%) |
| History of obstructive sleep apnea | 201 (67%) |
| **Sleep Hygiene Practices** |  |
| ***Home Environment*** |  |
| Unsafe at night | 26 (9%) |
| Uncomfortable physical environment | 39 (13%) |
| Uncomfortable temperature | 24 (8%) |
| Noise disturbances | 81 (27%) |
| Light disturbances | 27 (9%) |
| ***Bedtime Behaviors*** |  |
| Watch TV | 181 (60%) |
| Use computer/tablet/phone | 198 (66%) |
| Play video games | 43 (14%) |
| Eat meals or snacks | 88 (29%) |
| ***Composite Score***, non-standardized mean ± SD | 0.27 (0.16) |
| *Abbreviations: GED: General Educational Development; SD: standard deviation |  |

### Associations Between Sleep Hygiene, Sleep Quality, and Psychological Stress

Table 2 displays the associations of the individual and the composite score of sleep hygiene with sleep quality as well as psychological stress. Feeling unsafe at night (β=0.74, *p* < 0.001), noise disturbances (β=0.51, *p*=0.019), and eating meals or snacks at bedtime (β=1.00, *p*=0.049) were positively related to poor sleep quality. Being unsafe at night (β=0.90, *p* < 0.001), having an uncomfortable physical environment (β=0.73, *p* < 0.001) or temperature (β=0.77, *p* < 0.001), and eating at bedtime (β=1.22, *p*=0.004) were positively related to higher stress. A comparison of standardized coefficients across all individual factors revealed that feeling unsafe was most strongly associated with both poor sleep quality and psychological stress. Furthermore, a higher composite score (indicating poorer home environment and bedtime behaviors) was significantly related to both poor sleep quality (β=0.61, *p*=0.011) and elevated psychological stress (β=0.60, *p*=0.003).

**Table 2.**
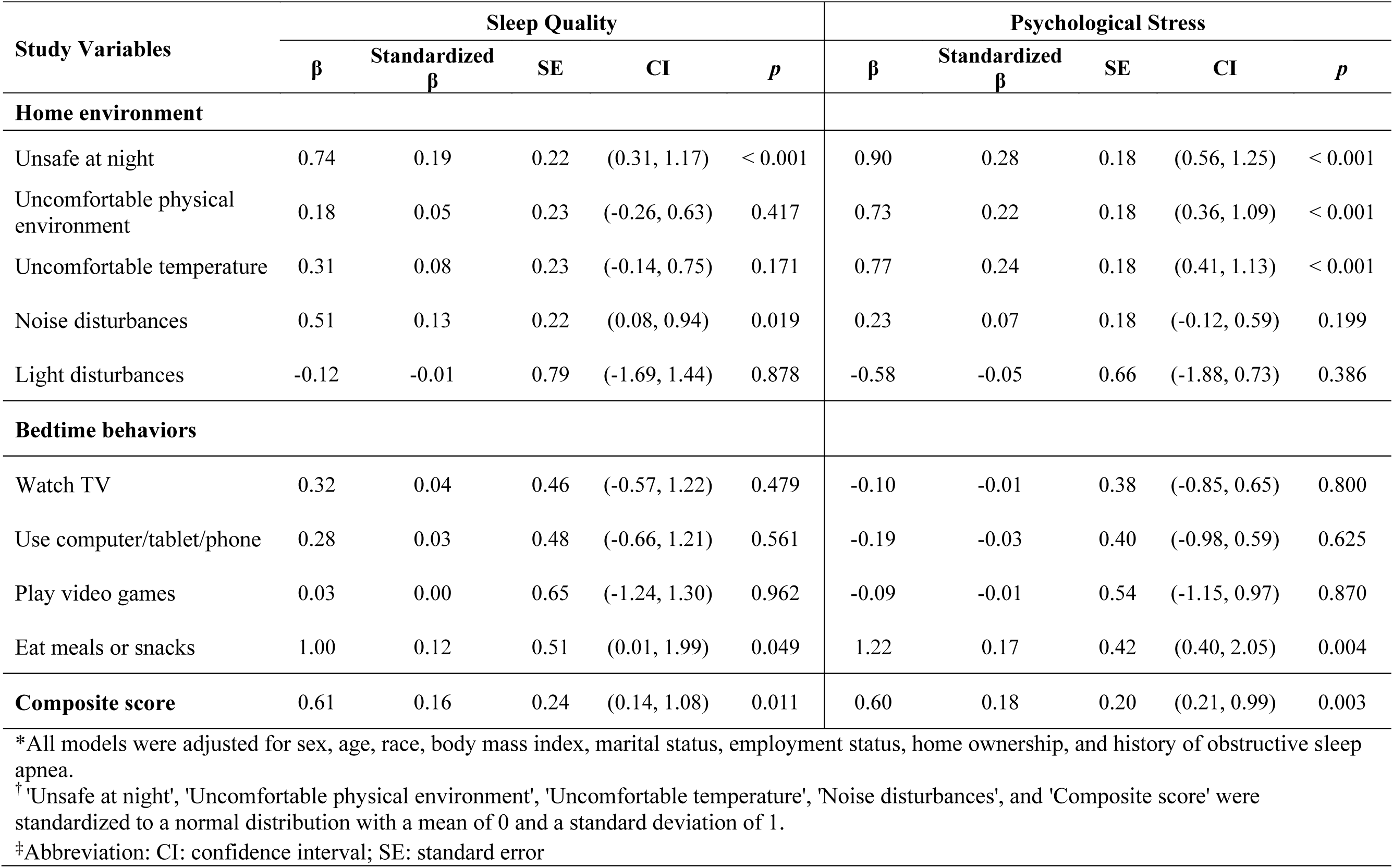
Associations Between Home Environment, Bedtime Behaviors, Sleep Quality, and Psychological Stress.

| Study Variables | Sleep Quality |  |  |  |  | Psychological Stress |  |  |  |  |
| --- | --- | --- | --- | --- | --- | --- | --- | --- | --- | --- |
| | $\beta$ | Standardized $\beta$ | SE | CI | *p* | $\beta$ | Standardized $\beta$ | SE | CI | *p* |
| **Home environment** |  |  |  |  |  |  |  |  |  |  |
| Unsafe at night | 0.74 | 0.19 | 0.22 | (0.31, 1.17) | < 0.001 | 0.90 | 0.28 | 0.18 | (0.56, 1.25) | < 0.001 |
| Uncomfortable physical environment | 0.18 | 0.05 | 0.23 | (-0.26, 0.63) | 0.417 | 0.73 | 0.22 | 0.18 | (0.36, 1.09) | < 0.001 |
| Uncomfortable temperature | 0.31 | 0.08 | 0.23 | (-0.14, 0.75) | 0.171 | 0.77 | 0.24 | 0.18 | (0.41, 1.13) | < 0.001 |
| Noise disturbances | 0.51 | 0.13 | 0.22 | (0.08, 0.94) | 0.019 | 0.23 | 0.07 | 0.18 | (-0.12, 0.59) | 0.199 |
| Light disturbances | -0.12 | -0.01 | 0.79 | (-1.69, 1.44) | 0.878 | -0.58 | -0.05 | 0.66 | (-1.88, 0.73) | 0.386 |
| **Bedtime behaviors** |  |  |  |  |  |  |  |  |  |  |
| Watch TV | 0.32 | 0.04 | 0.46 | (-0.57, 1.22) | 0.479 | -0.10 | -0.01 | 0.38 | (-0.85, 0.65) | 0.800 |
| Use computer/tablet/phone | 0.28 | 0.03 | 0.48 | (-0.66, 1.21) | 0.561 | -0.19 | -0.03 | 0.40 | (-0.98, 0.59) | 0.625 |
| Play video games | 0.03 | 0.00 | 0.65 | (-1.24, 1.30) | 0.962 | -0.09 | -0.01 | 0.54 | (-1.15, 0.97) | 0.870 |
| Eat meals or snacks | 1.00 | 0.12 | 0.51 | (0.01, 1.99) | 0.049 | 1.22 | 0.17 | 0.42 | (0.40, 2.05) | 0.004 |
| **Composite score** | 0.61 | 0.16 | 0.24 | (0.14, 1.08) | 0.011 | 0.60 | 0.18 | 0.20 | (0.21, 0.99) | 0.003 |

\*All models were adjusted for sex, age, race, body mass index, marital status, employment status, home ownership, and history of obstructive sleep apnea.
† 'Unsafe at night', 'Uncomfortable physical environment', 'Uncomfortable temperature', 'Noise disturbances', and 'Composite score' were standardized to a normal distribution with a mean of 0 and a standard deviation of 1.
‡Abbreviation: CI: confidence interval; SE: standard error

### Mediation Analysis of Sleep Hygiene, Sleep Quality, and Psychological Stress

Results from the mediation analysis showed that the composite score of sleep hygiene was significantly associated with sleep quality (path a: β=0.61, standard error [SE] = 0.25, *p*=0.014) and that sleep quality was significantly associated with psychological stress while adjusting for the composite score (path b: β=0.27, SE = 0.05, *p* < 0.001). After adjusting for sleep quality, the direct relationship between the composite score and psychological stress remained statistically significant (β=0.44, SE=0.173, *p* =0.012). The indirect effect, assessed using a bias-corrected bootstrap confidence interval, indicated that sleep quality partially mediated the relationship between the composite score and psychological stress (β=0.16, 95% BCa CI:0.04-0.35). Sleep quality accounted for approximately 27% of the total association between the composite score and psychological stress (Figure 2).

**Figure 2.**
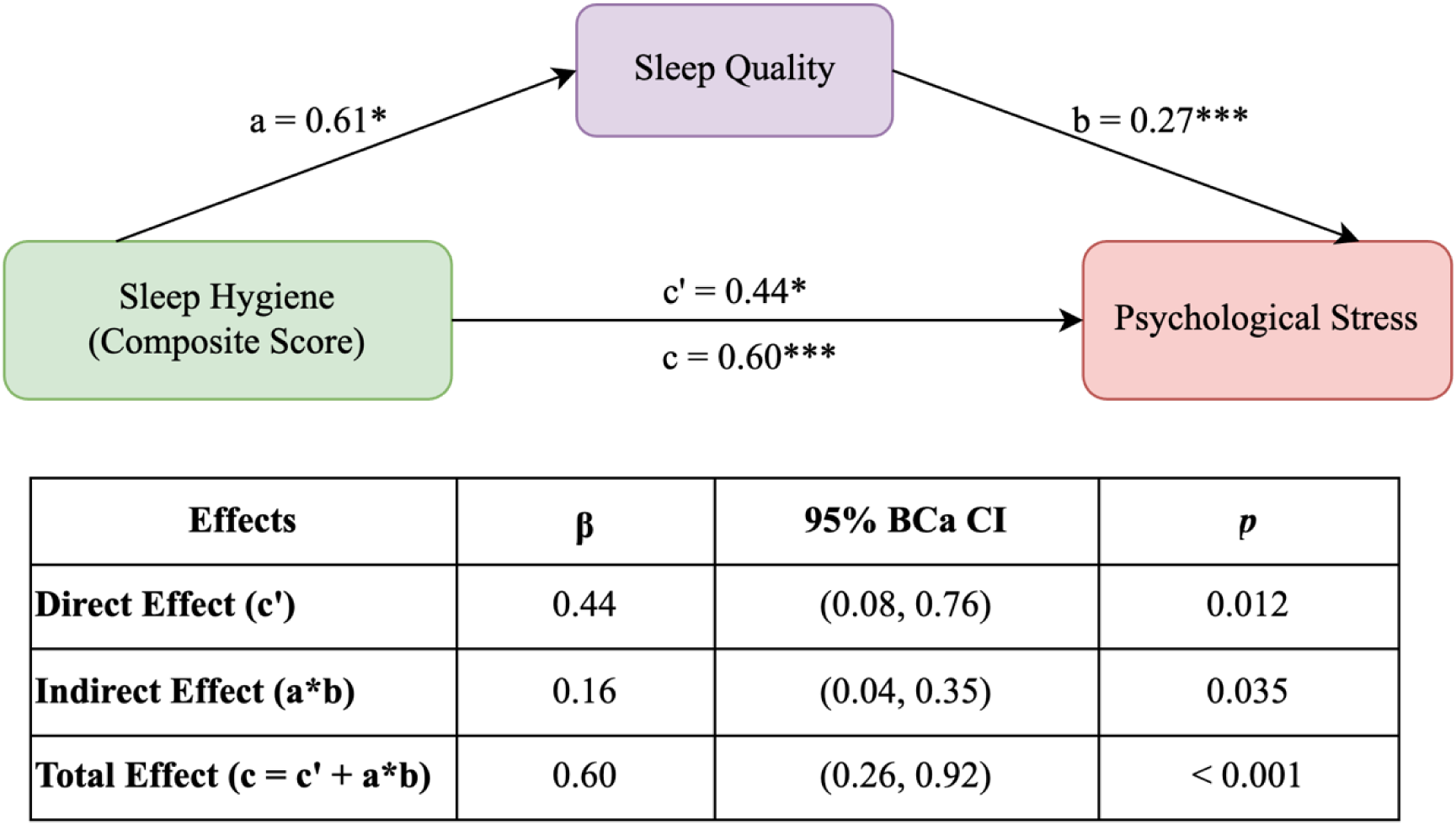
Sleep quality mediates the association between the composite score of sleep hygiene and psychological stress. a, b, c, and c’ denote path coefficients. The c path coefficient represents the total effect of sleep hygiene composite score on psychological stress. The c’ path coefficient represents the direct effect of sleep hygiene composite score on psychological stress after controlling for sleep quality. Analyses were adjusted for sex, age, race, body mass index, marital status, employment status, home ownership, and history of obstructive sleep apnea. *\*\*\*p* < 0.001, *\*\* p* < 0.01, *\*p* < 0.05 Abbreviation: BCaCI: bias-corrected and accelerated bootstrap confidence interval

## DISCUSSION

Our study examined the relationship between room environment, bedtime behaviors, sleep quality, and psychological stress in a cohort of 300 adults diagnosed with hypertension and diabetes. Sleep hygiene practices, including room environment and bedtime behaviors, played crucial roles in sleep and mental well-being. In addition, we discovered that poor room environment and bedtime behaviors may exacerbate psychological stress through poor sleep quality. While the causal relationship remains to be established, our findings underscore the critical need to promote healthy sleep hygiene. Doing so may significantly enhance sleep quality and reduce stress in adults with multiple CVD risk factors.

A key finding of our study is that 78% of adults with multiple CVD risk factors experienced poor sleep quality. CVD risk factors can disrupt sleep physiology which consequently leads to sleep disturbances and fragmentation. This disruption may result from mechanisms such as sympathetic hyperactivity due to elevated blood pressure,^28^ peripheral neuropathy,^29^ and physical discomfort^30^ (e.g., nocturia) caused by unstable blood sugar levels. In particular, the co-occurrence of hypertension and diabetes increases the risk of OSA, which was observed in 67% of this study cohort, and is another key determinant affecting sleep. Together, all these factors likely explain the high prevalence of poor sleep quality among our participants. Without proper management, poor sleep aggravates CVD risk factors and leads to series of complications especially when the risk factors overlap – as demonstrated by elevated psychological stress in our sample. The finding aligns with data from a recent national survey, which revealed that 46% of U.S. adults (N = 1,000) with below-average sleep quality rated their mental health as poor or very poor.^31^ Therefore, improving sleep quality is crucial to break this cycle and reduce CVD risks.

This study revealed suboptimal room environment and unhealthy bedtime behaviors among adults with multiple CVD risk factors. Noise disturbances were the most commonly reported adverse environmental factor. Additionally, the findings highlighted the prevalent use of electronic devices in people’s bedtime routines, with 66% of participants using small screens and 60% watching TV in bed. These behaviors, though often normalized, are known to increase alertness by delaying melatonin release and disrupting the body’s natural sleep-wake cycle. Addressing the aforementioned issues through targeted interventions such as fostering awareness about healthy sleep hygiene could play a pivotal role in improving the cardiovascular and overall health of individuals with these risk factors.

Consistent with previous evidence^32^, our study identified specific factors influencing sleep quality and psychological stress. Noise disturbances were strongly linked to poor sleep quality, while an uncomfortable environment and temperature were associated with elevated stress. We also identified shared factors that simultaneously affected sleep quality and psychological stress. In particular, nighttime safety concerns demonstrated the strongest relationship with both, underscoring home safety as a vital social determinant of health in shaping individual well-being. Other researchers have similarly pointed out the connection between safety and health outcomes. A study of participants living in low socioeconomic status showed that individuals who reported fears related to safety during sleep were more likely to report poor quality of sleep, as indicated by a PSQI score over 5, in comparison to those without such concerns.^33^ Adults residing in low-income homes in New York City expressed concerns about home security increased their stress levels.^32^ Another shared factor influencing both sleep quality and psychological stress was consuming food at bedtime. Data from the American Time Use Survey showed a linkage between eating or drinking < 1 hour before bedtime and increased wake after sleep onset, an objective sleep quality metric.^34^ Chronic stress can promote seeking and intake of high-fat and energy-dense foods through increased production of cortisol, and for some individuals, eating meals or snacks at bedtime may function as a coping mechanism for alleviating psychological stress.^35^ However, such behaviors can inadvertently worsen sleep quality, creating a feedback loop of poor sleep and heightened stress. One should note that the composite score was strongly related to both poorer sleep quality and elevated stress. The finding holds important public health and clinical implications because addressing bedroom environment and bedtime behaviors may be effective in achieving optimal health outcomes. They align with prior research, such as Johnson et al.,^17^ which highlighted the role of household conditions and in-bed behaviors in contributing to sleep disturbances. However, our study advances the understanding of these interactions by demonstrating their combined effects on psychological stress.

We additionally found that sleep quality partially mediated the relationship between sleep hygiene and stress. Although a causal relationship cannot be inferred, the mediation analysis suggests that poor sleep hygiene is linked to stress both directly and indirectly through its impact on sleep quality. Therefore, promoting sleep hygiene practices may be an efficient, cost-effective approach to addressing sleep and mental health issues, though this approach requires further testing to confirm its effectiveness. While existing sleep interventions vary in format^36^, we posit that delivering education on healthy sleep hygiene practices may yield potential health benefits. This is supported by several studies. A randomized controlled trial (RCT) conducted among shift work nurses found that those who received a 2-hour training on sleep hygiene practices showed significantly lower sleep latency, better perceived sleep quality, and lower insomnia and daytime sleepiness symptoms in comparison to the control group.^37^ Another RCT by Kloss et al.^38^ observed improved sleep hygiene knowledge, fewer maladaptive beliefs and attitudes about sleep, and shorter sleep onset in college students who participated in two 90-minute workshops on sleep hygiene practices. Future research should aim to explore the most effective strategies for improving sleep and reducing stress.

There are several limitations of our study that should be noted. As aforementioned, the cross-sectional nature of this study prevents us from inferring causal relationships between the variables studied. Longitudinal studies are needed to confirm these findings. Second, our participants were comprised of adults receiving primary care within a single healthcare system in Maryland, which may limit the generalization of the results to the broader U.S. population. Third, while our study assessed important aspects of sleep hygiene practices, it did not capture all relevant factors. Other important practices, such as maintaining consistent sleep schedules, limiting the intake of stimulating foods (e.g., alcohol, caffeine), exercising regularly, and avoiding naps in the late afternoon or evening, were not included in our evaluation.^39^ Lastly, sleep was self-reported in this study. Although the PSQI provides a reliable measure of perceived sleep quality, self-reporting is often subject to recall bias and may not fully align with objective measures.

In conclusion, this study identified strong associations between sleep hygiene practices, sleep quality, and stress in adults with multiple CVD risk factors. Specifically, both individual sleep hygiene components—household environment and bedtime behaviors—and the composite score were independently linked to poor sleep quality and elevated stress. The findings suggest a potential pathway in which better sleep hygiene practices enhance sleep quality, which could potentially reduce psychological stress. Given the crucial roles of both sleep and stress in cardiovascular health, promoting modifiable sleep hygiene practices may be an effective strategy to mitigate health risks in this population.

## Data Availability

The data will not be available.

## ACKNOWLEDGMENTS

None

